# Direct estimates of absolute ventilation in primary health care clinics in South Africa

**DOI:** 10.1101/2022.03.17.22272421

**Authors:** Peter G Beckwith, Aaron S Karat, Indira Govender, Arminder Deol, Nicky McCreesh, Karina Kielmann, Kathy Baisley, Alison D Grant, Tom A Yates

**Author notes:** **Corresponding author** Dr Peter Beckwith, Department of Clinical Research, Faculty of Infectious and Tropical Diseases, London School of Hygiene & Tropical Medicine, Keppel Street, London WC1E 7HT, United Kingdom |.

## Abstract

**Background:** Healthcare facilities are important sites for the transmission of pathogens spread via bioaerosols, such as *Mycobacterium tuberculosis* (*Mtb*). Natural ventilation can play an important role in reducing this transmission. In primary health care (PHC) clinics in low and middle-income settings, susceptible people, including healthcare workers, are exposed to individuals with infectious pulmonary tuberculosis. We measured rates of natural ventilation in PHC clinics in KwaZulu-Natal and Western Cape provinces, South Africa.

**Methods and Findings:** We measured ventilation in clinic spaces using a tracer-gas release method. In spaces where this was not possible, we estimated ventilation using data on indoor and outdoor carbon dioxide levels, under reasonable assumptions about occupants’ metabolic rates. Ventilation was measured i) under usual conditions and ii) with all windows and doors fully open. We used these ventilation rates to estimate the risk of *Mtb* transmission using the Wells-Riley Equation. We obtained ventilation measurements in 33 clinical spaces in 10 clinics: 13 consultation rooms, 16 waiting areas and 4 other clinical spaces. Under usual conditions, the absolute ventilation rate was much higher in waiting rooms (median 1769 m^3^/hr, range 338–4815 m^3^/hr) than in consultation rooms (median 197 m^3^/hr, range 0–1451 m^3^/hr). Ventilation was better in permanent than in temporary structures. When compared with usual conditions, fully opening existing doors and windows resulted in a median two-fold increase in ventilation. Our Wells-Riley estimates show that, following sustained exposure, or contact with highly infectious index cases, some risk of *Mtb* infection may persist in the best ventilated clinical spaces unless other components of transmission risk are also addressed.

**Conclusions:** Among the clinical spaces studied, we observed substantial variation in natural ventilation. Ventilation interventions may have considerable impact on *Mtb* transmission in this setting. We recommend these form part of a package of infection prevention and control interventions.

## Introduction

Good ventilation in congregate spaces can reduce transmission of airborne pathogens, such as *Mycobacterium tuberculosis* (*Mtb*), Rubeola virus (measles), and SARS-CoV-2[1] [2][3]. However, ventilation rates are difficult to measure and there are limited data available on ventilation rates in public spaces, such as clinics, in sub-Saharan Africa [4][5][6][7][8][9].

Healthcare facilities bring together infectious and susceptible individuals and are important sites for the transmission of airborne pathogens. This is particularly true for *Mtb* [10]. Clinic attendees including vulnerable populations, such as people living with HIV, may inhale bioaerosols containing *Mtb* produced by individuals with pulmonary tuberculosis (TB) attending the same facility [11][10][12][13][14]. *Mtb* is an important occupational health concern, with health workers at greater risk than the general population of infection and reinfection and, as a result, developing active TB disease [15][16].

Improved ventilation, an infection prevention and control (IPC) intervention, decreases the risk of infection by removing droplet nuclei containing aerosolised *Mtb*. Natural ventilation plays a key role in low and middle-income settings where the resources and infrastructure needed for mechanical ventilation systems are usually unavailable [17]. Natural ventilation can have comparable or superior performance to mechanical ventilation systems [17]. However, rates will vary with changes in wind speed or direction, and inadequate rates of natural ventilation may occur in poorly designed buildings [18].

We aimed to describe the ventilation of waiting areas, consultation rooms, and other clinical spaces across ten primary healthcare clinics (PHCs) in South Africa. Our ventilation experiments were undertaken as part of an interdisciplinary project called *Umoya omuhle* (meaning “good air” in Zulu), which used a whole systems approach to understand *Mtb* transmission and TB IPC in PHC clinics in South Africa [19][20].

## Methods

### Setting

Ventilation experiments were performed in five PHC clinics in KwaZulu-Natal and five in Western Cape province, South Africa, between December 2018 and December 2019. These facilities were built between the 1980s and early 2010s and serve both urban and rural populations. They were selected to be broadly representative of PHC clinics in the two provinces with respect to location, age of building, and type of clinic. Each clinic had a unique design, though some clinics had several rooms that were identical in their layout. Many clinics had a mixture of permanent and temporary buildings. Temporary buildings included “park homes” (portacabins) that were used to rapidly expand clinic capacity as the antiretroviral therapy programme was being rolled out in the 2000’s [21].

Measurements were undertaken in spaces where patients either waited or interacted with clinic staff. These included: rooms where patient vital signs were measured; rooms where phlebotomy was performed; formal and informal waiting areas; and consultation rooms. The timing of experiments was agreed in advance with clinic managers.

### Weather

Data on wind speed and temperature were provided by the South African Weather Services for the period 01 January 2018 to 31 December 2020. These consisted of hourly measurements taken at the nearest weather station to each clinic (median Euclidean distance: 20.5km [range: 5-35km]). Using these data, the wind speed and temperature at the time experiments were undertaken were compared to the distribution observed during typical clinic opening hours (0600–1800) across this three-year period.

### Describing spaces

The dimensions of each room were measured using a Bosch PLR 40R digital laser measure (Bosch, Gerlingen, Germany; accuracy +/-2 mm) and room volumes calculated. The configuration of the windows and doors when the room was in routine use was recorded – these were then used as the‘usual’ conditions in our experiments. To summarise these data, window configuration was classified into one of four categories: 1) open (all windows fully open); 2) closed (all windows closed); 3) half or less open; or 4) more than half open. For example, if there were three windows, one closed and two half open, the windows would be 33% open [(0 + 0.5 + 0.5) / 3], so the windows would be classified as ‘half or less open’. The same classification was used for doors.

### Measuring ventilation

#### Tracer gas release experiments

Our primary means of measuring ventilation was a tracer gas release technique using carbon dioxide (CO_2_; supplementary table 1) [17][22]. Ventilation was estimated under both ‘usual’ conditions (the configuration observed when the room was in routine use) and ‘ideal’ conditions (with existing windows and doors maximally open; supplementary table 2).

At the start of each experiment, every window and door was closed and any remaining openings were taped closed with plastic sheeting. CO_2_ monitors were placed at two different central locations within the room (Datalogging Indoor Air Quality Meter – 800050, Sper Scientific, Scottsdale, Arizona, USA; accuracy +/-75 parts per million [ppm] CO_2_). The concentration of CO_2_ in the room was then increased by releasing CO_2_ fire extinguishers for 5–10 seconds. CO_2_ was mixed with room air using a paddle fan for two minutes, with the aim of achieving a stable and homogeneous CO_2_ concentration of 3,000–10,000 ppm throughout the space. The fan was then turned off and, after five minutes, any plastic sheeting was removed and the windows and doors were placed in either the usual or ideal configuration. CO_2_ levels were then recorded every second for a period of approximately five minutes. The aim was to perform three experiments under usual conditions, and three under ideal conditions, yielding six CO_2_ decay curves under each set of conditions.

#### Rebreathed fraction approach

Tracer gas release experiments were not possible in spaces that 1) could not be vacated by patients or clinic staff or 2) were large and very open to the outdoors, meaning high levels of CO_2_ could not be attained at baseline. Where tracer gas release experiments were not possible, an established approach [23] [24] was adapted to estimate the ventilation rate using data on indoor CO_2_ concentrations, outdoor CO_2_ concentrations, and occupancy [25]. These experiments were undertaken in eight main waiting areas during working hours when spaces were occupied by patients and staff.

In these waiting areas, three CO_2_ monitors were placed at different central locations and one monitor placed immediately outside the building to capture the CO_2_ concentration of the replacement air. Measurements were obtained every second. The number of individuals in the room was counted every ten minutes, categorising individuals as aged <1 year, 1–5 years, or >5 years. Data were collected for between one and three hours under each set of conditions (usual and ideal).

### Statistical analysis

The tracer gas release experiments produced data similar to those presented in Supplementary Figure 1. Analysis focussed on the right-hand side of these curves. Specifically, data were included from 30 seconds after the doors and windows were opened until CO_2_ levels reached 200 ppm above baseline or, where no baseline value was available, until CO_2_ levels reached 800 ppm.

Prior to analysis, the CO_2_ data were smoothed, using a sixty second moving average (mean), consisting of the 30 seconds before and after each measurement. Using these smoothed data, the natural logarithms of the CO_2_ concentration measurements were plotted against time in hours. A linear regression line was then fitted through each trace, with the gradient of this line providing an estimate of the number of air changes per hour (ACH). With (typically) six such estimates per experiment (three experiments, two CO_2_ meters per experiment), a pooled estimate of the number of air changes per hour, with an associated 95% confidence interval (CI), was obtained using random effects meta-analysis. The absolute ventilation rate (in m^3^/hour) was then calculated as the product of the number of air changes per hour and the room volume.

To analyse the paired indoor and outdoor CO_2_ measurements, we developed an adaptation of the Persily and de Jonge method [24] to allow for non-steady state conditions. The outside CO_2_ concentration was estimated as a mean obtained through curve fitting, using linear interpolation. The indoor CO_2_ concentration in each space was calculated at each time point by taking the mean value from the three inside monitors. The number of individuals in each age group in the room at each time point was multiplied by their assumed CO_2_ generation rate to estimate the total CO_2_ generation rate at each time point. Age-specific CO_2_ production rates were taken from Persily and de Jonge [24], assuming an activity rate of 1.2 MET (occupants sitting quietly). A simple linear regression model was then fitted describing how, at each time point, the difference between the indoor CO_2_ concentration and the outdoor CO_2_ concentration related to the total CO_2_ generation rate. The slope of this regression line estimated the absolute ventilation rate. More details of this method are given elsewhere[25]. Note, outdoor CO2 concentrations in our experiments were measured and found to be stable. In settings where outdoor CO2 levels are more variable, investigators using this method may prefer to update the outdoor CO2 measurement at each timestep.

Most spaces were only visited on one occasion and, as such, our estimates of ventilation do not fully capture variation in ventilation rates associated with, for example, changes in wind speed or direction. For this reason, when combining data from different spaces, we opted for a descriptive presentation of our results. We avoided calculating pooled effect estimates with associated confidence intervals, as we did not wish to overestimate precision.

### Calculation of risk

The Wells-Riley equation models transmission risk for airborne infections as a Poisson process. The approach assumes a well-mixed airspace. It takes into account the time spent in a space, the number of infectious individuals present, the number of ‘infectious quanta’ they produce per unit time (usually assumed), the volume of air susceptible individuals breathe per unit time, and the absolute ventilation rate [26]. Here, quanta are defined as ‘the number of infectious airborne particles required to infect which may be one or more airborne particles’ [26].

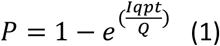

P = probability for each susceptible individual of being infected over time t; I = Number of infectious individuals; q = number of infectious quanta they produce per hour; p = pulmonary ventilation rate of susceptible individuals (volume/time); t = exposure time; Q = absolute room ventilation (volume/time)

We related our calculated absolute ventilation rate to the risk of *Mtb* transmission using the Wells Riley Equation[26]. These calculations assumed a single infectious individual was present and that susceptible individuals had a pulmonary ventilation rate of 0.6 m^3^/hr.

The infectiousness of people with pulmonary TB is known to be heterogeneous. The standard assumption is that infectious individuals produce 1.25 infectious quanta per hour, based on data from early studies at Johns Hopkins [27]. A similar study of HIV-positive adults with pulmonary TB in Lima, Peru, reported a mean quanta production rate of 8.2 per hour, and the most infectious individual in that study produced 226 quanta per hour [28]. We, therefore, estimated transmission risk at 1.25, 8.2 and 226 quanta/hr.

### Tools and Code

Analysis of the tracer gas release experiments was undertaken in Stata version 14.2 (StataCorp, College Station, Texas, USA). This Stata code is available on GitHub (https://github.com/tayates/uo_ventilation). Analysis of paired indoor-outdoor CO_2_ measurements was carried out using R version 3.6.0 [29].This R code is also available on GitHub (URL: https://github.com/ArminderD/ventilation.git). The data are available via https://datacompass.lshtm.ac.uk/.

### Ethics

The *Umoya omuhle* study received ethical approval from the Biomedical Research Ethics Committee of the University of KwaZulu-Natal (ref. BE082/18), the Human Research Ethics Committee of the Faculty of Health Sciences of the University of Cape Town (ref. 165/2018), the Research Ethics Committee of Queen Margaret University (ref. REP 0233), and the Observational/Interventions Research Ethics Committee of the London School of Hygiene & Tropical Medicine (ref. 14872). No data on individuals were captured for this aspect of the project. Consent to undertake these experiments was obtained from facility managers.

## Results

### Setting

Ventilation experiments were performed in 33 clinic spaces between December 2018 and December 2019. Clinic characteristics are detailed in Table 1. Tracer release experiments were performed in 26 different clinic spaces of which 13 (50%) were consulting rooms; nine (35%) were waiting areas; and four (15%) other spaces, including corridors, rooms where patients’ vital signs were measured, and phlebotomy rooms. Eight main waiting areas were assessed using the rebreathed fraction approach.

**Table 1:**
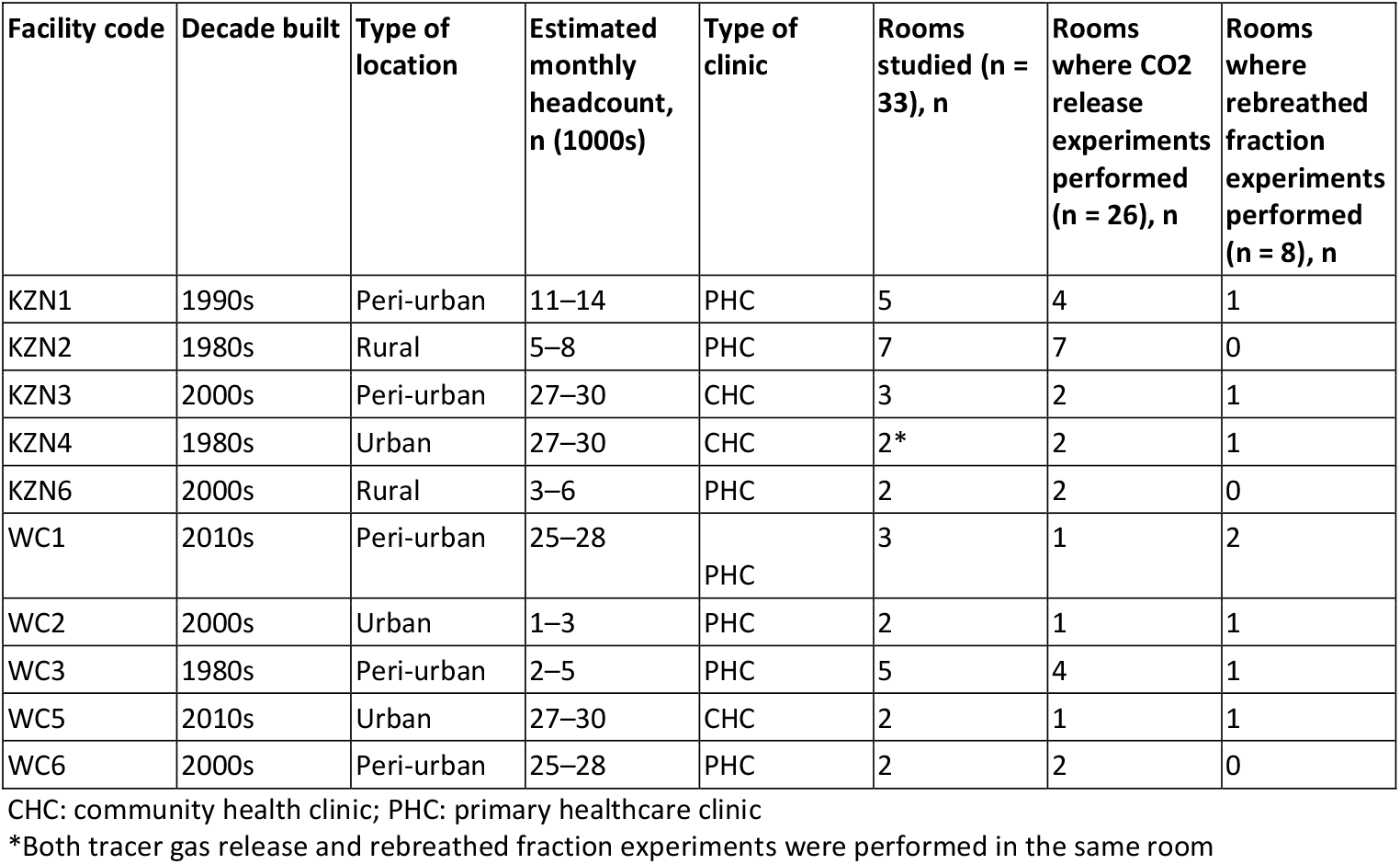
Characteristics of facilities (n = 10) and rooms (n = 33) in which ventilation experiments were conducted

| Facility code | Decade built | Type of location | Estimated monthly headcount, n (1000s) | Type of clinic | Rooms studied (n = 33), n | Rooms where CO<sub>2</sub> release experiments performed (n = 26), n | Rooms where rebreathed fraction experiments performed (n = 8), n |
| --- | --- | --- | --- | --- | --- | --- | --- |
| KZN1 | 1990s | Peri-urban | 11–14 | PHC | 5 | 4 | 1 |
| KZN2 | 1980s | Rural | 5–8 | PHC | 7 | 7 | 0 |
| KZN3 | 2000s | Peri-urban | 27–30 | CHC | 3 | 2 | 1 |
| KZN4 | 1980s | Urban | 27–30 | CHC | 2* | 2 | 1 |
| KZN6 | 2000s | Rural | 3–6 | PHC | 2 | 2 | 0 |
| WC1 | 2010s | Peri-urban | 25–28 | PHC | 3 | 1 | 2 |
| WC2 | 2000s | Urban | 1–3 | PHC | 2 | 1 | 1 |
| WC3 | 1980s | Peri-urban | 2–5 | PHC | 5 | 4 | 1 |
| WC5 | 2010s | Urban | 27–30 | CHC | 2 | 1 | 1 |
| WC6 | 2000s | Peri-urban | 25–28 | PHC | 2 | 2 | 0 |

CHC: community health clinic; PHC: primary healthcare clinic
\*Both tracer gas release and rebreathed fraction experiments were performed in the same room

Under usual conditions in the 26 rooms where the CO_2_ release technique was used, four rooms (15%) had no windows, two (8%) had all existing windows closed, 12 (46%) had their windows less than half open, and eight (31%) had their windows maximally open (of which the majority were consulting rooms (5/8 [63%]). All consulting rooms had their doors closed when attending to patients and no waiting rooms had more than half of their doors open when the room was in use (Table 2).

**Table 2:** The configuration of windows and doors under usual conditions in rooms where tracer gas release experiments were performed (n = 26 experiments on 12 days)

| Type of room | Configuration of windows |  |  |  | Configuration of door in rooms with one door (n = 13), n |  | Configuration of doors in rooms with multiple doors (n = 13), n |  |  |
| --- | --- | --- | --- | --- | --- | --- | --- | --- | --- |
|  | No windows | All windows closed | Windows open half or less | All windows maximally open | Door closed | Door open | All doors closed | Half or less doors open | Greater than half doors open |
| Consulting room (n = 13) | 0 | 2 | 6 | 5 | 11 | 0 | 0 | 1 | 1 |
| Waiting area (n = 9) | 3 | 0 | 4 | 2 | 0 | 0 | 0 | 7 | 2 |
| Other (n = 4)* | 1 | 0 | 2 | 1 | 1 | 1 | 0 | 0 | 2 |
| **Total (n = 26)** | **4** | **2** | **12** | **8** | **12** | **1** | **0** | **8** | **5** |

\*Other rooms included: two rooms where patient vital signs were measured, one phlebotomy room, one corridor where patients waited

### Temperature and wind speed

We compared temperature and wind speed measured at the nearest weather station at the time we undertook our experiments to the mean temperature and wind speed measured during typical clinic opening hours at the same weather station.

Temperatures at the times tracer gas release experiments were conducted were within one and two standard deviations (SDs) of the 3-year mean temperature for 10/12 (83%) and 12/12 (100%) experiment days, respectively, and wind speeds within one and two SDs of the 3-year mean wind speed for 10/12 (83%) and 11/12 (92%) experiment days, respectively (Figure 1).

**Figure 1.**
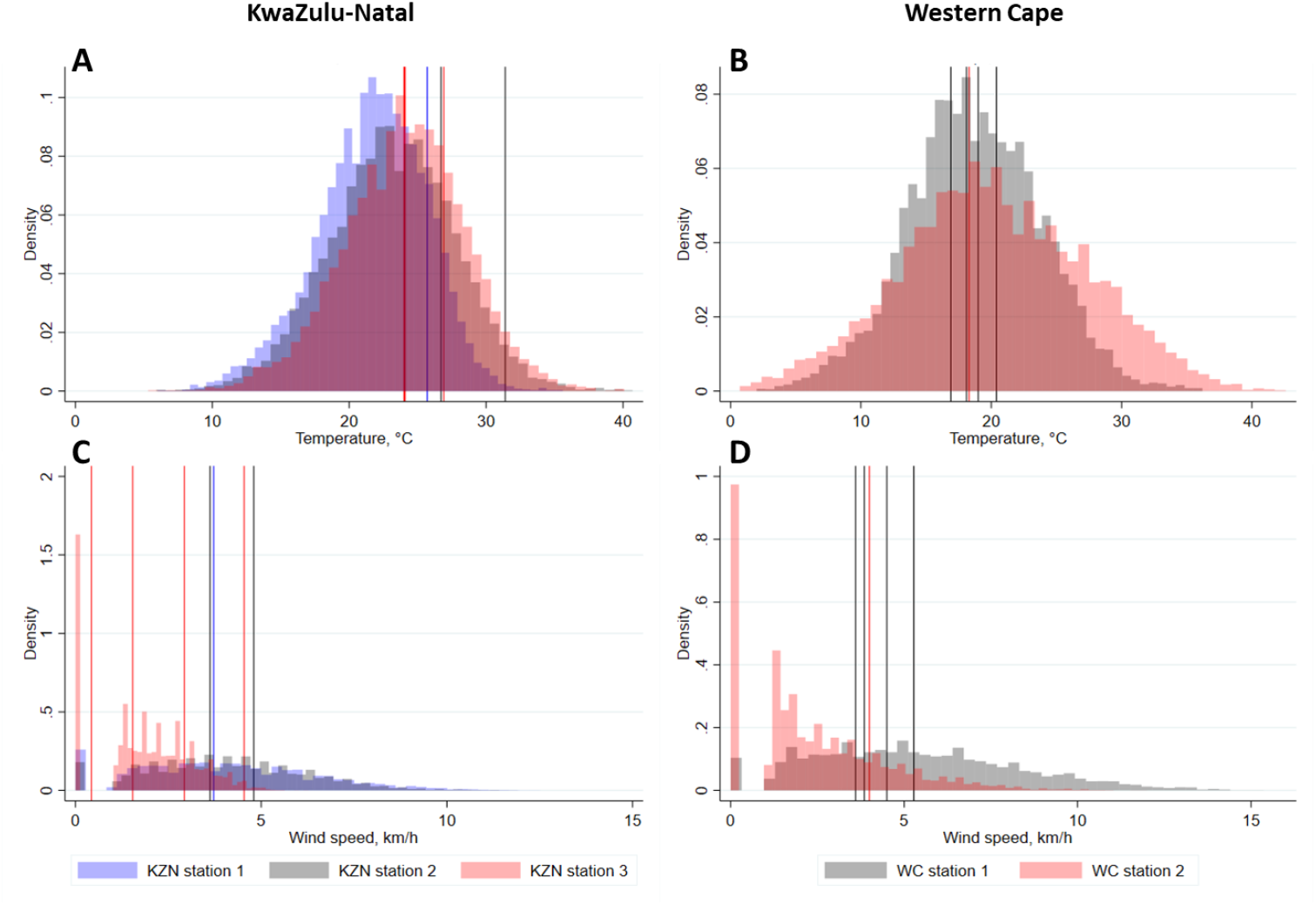
Histograms showing the distribution of temperatures and wind speeds in KwaZulu-Natal and Western Cape during working hours from January 2018–December 2020. Vertical lines show the mean temperatures and wind speeds on the 12 days when tracer gas release experiments were conducted A: temperatures in KwaZulu-Natal; B: temperatures in Western Cape; C: wind speeds in KwaZulu-Natal; D: wind speeds in Western Cape. Vertical lines indicate mean temperature or wind speed at the weather station closest to each clinic during working hours on the day the data were collected. Colour of histograms and vertical lines corresponds to specific weather stations in each province. C: centigrade; km/h: kilometres per hour; KZN: KwaZulu-Natal; WC: Western Cape

Temperatures at the time we performed rebreathed fraction experiments were within one and two SDs of the 3-year mean temperature for 7/8 (88%) and 8/8 (100%) experiment days and wind speeds within one and two SDs of the 3-year mean wind speed for 7/8 (88%) and 8/8 (100%) of experiment days (Supplementary Figure 2).

### Tracer gas release experiments

The distribution of room volumes across the 26 spaces in which we performed tracer gas release experiments is presented in Figure 2A. Box and whisker plots in Figure 3 present the same results disaggregated by province, type of room, the age of the building, and whether the community is urban or rural. The distribution of room sizes was skewed with a small number of much larger rooms. As might be expected, consultation rooms (n = 13; median volume 31 m^3^, range 15–45 m^3^) were smaller than waiting rooms (n = 9; median volume 68m^3^, range 31–135 m^3^). Rooms in temporary structures (n = 5; median volume 21 m^3^, range 15–106 m^3^) were smaller than those in permanent structures (n = 21; median volume 43 m^3^, range 21–135 m^3^).

**Figure 2:**
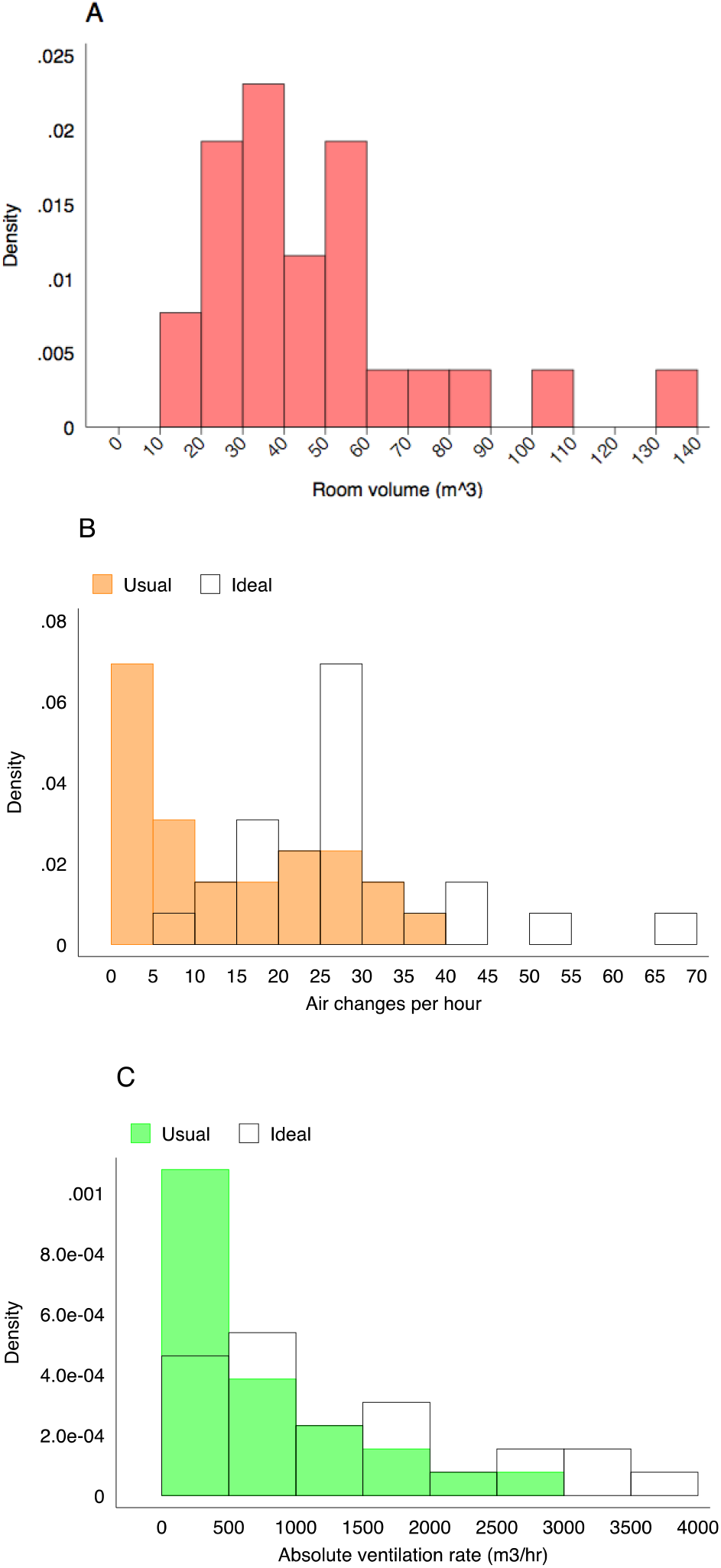
Histograms describing the distribution of room volumes (A), the number of air changes per hour (B) and the absolute ventilation rate (C) in the 26 clinical spaces where we conducted tracer gas release experiments. Ventilation rates are described under usual conditions and ideal conditions (all doors and windows fully open)

**Figure 3:**
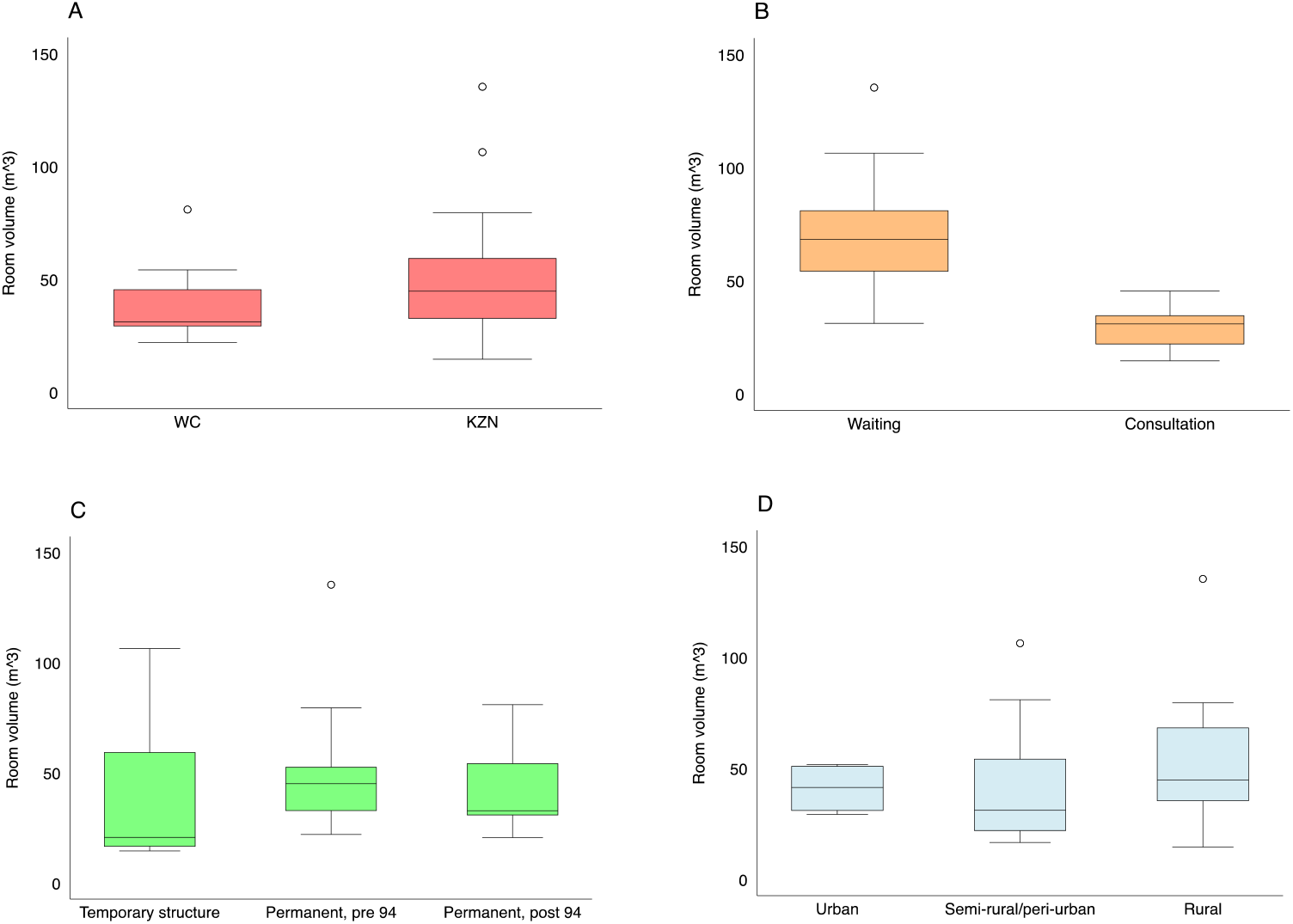
Box and whisker plots describing the distribution of room volumes in the clinical spaces where we conducted tracer gas release experiments. Results are disaggregated by province (A), room type (B), the age of the building (C), and clinic location (D). Here, the boxes mark the 25^th^ percentile, the 50^th^ percentile (median) and the 75^th^ percentile, with the whiskers marking the upper and lower adjacent values.

The distribution of the number of ACH under both usual and ideal conditions is presented in Figure 2B. The same data disaggregated by province, type of room, the age of the building, and whether the community is urban or rural are presented in Figure 4. The number of ACH varied substantially. It was higher in waiting rooms than in consultation rooms (usual conditions: median ACH in waiting rooms = 21, range 4 – 36; median ACH in consultation rooms = 4, range 0 - 32) and, as expected, higher with existing doors and windows fully open (ideal conditions: median ACH in waiting rooms = 29, range 16 - 65; median ACH in consultation rooms = 26, range 7 - 54). Opening existing doors and windows resulted in a median 2.1-fold increase in the number of air changes per hour (range 1 - 25.6; note, this range does not include a consultation room with zero effective ventilation under usual conditions).

**Figure 4:**
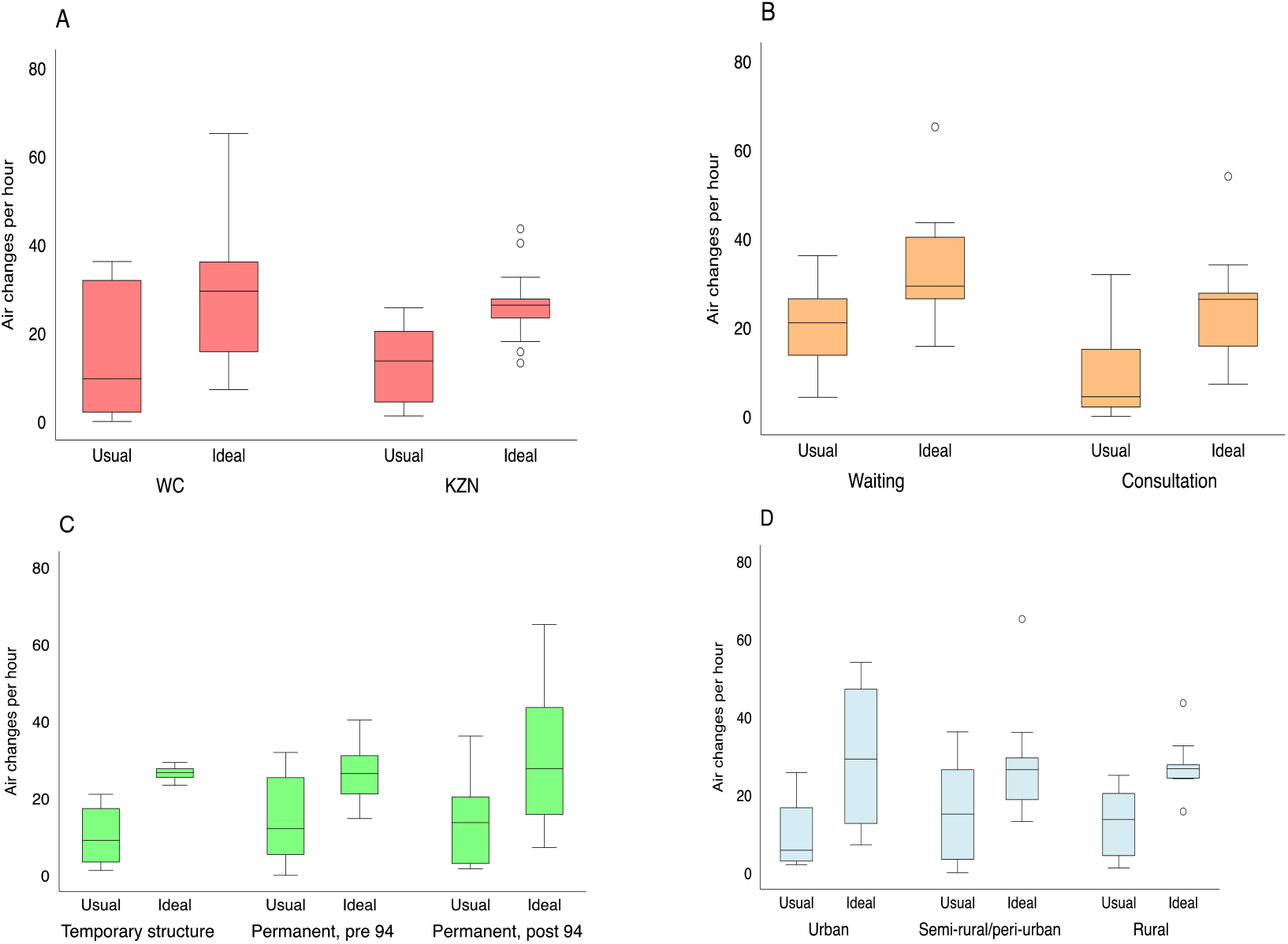
Box and whisker plots describing the distribution of the number of air changes per hour in the 26 clinical spaces where we conducted tracer gas release experiments. Ventilation rates are described under usual conditions and with all doors and windows fully open. Results are disaggregated by province (A), room type (B), the age of the building (C), and clinic location (D). Here, the boxes mark the 25^th^ percentile, the 50^th^ percentile (median) and the 75^th^ percentile, with the whiskers marking the upper and lower adjacent values.

The distribution of the absolute ventilation rates is presented in Figure 2C. These data, disaggregated by province, type of room, the age of the building, and whether the community is urban or rural, are presented in Figure 5. Again, we observed substantial variation. The absolute ventilation rate was much higher in waiting rooms (usual conditions: median absolute ventilation rate = 1289 m^3^/hr, range 338–2651 m^3^/hr) than in consultation rooms (usual conditions: median absolute ventilation rate 197 m^3^/hr, range 0–1451 m^3^/hr). Temporary buildings (usual conditions: median absolute ventilation rate 360 m^3^/hr, range 18–2240 m^3^/hr) were less well ventilated than permanent structures (usual conditions: median absolute ventilation rate 493 m^3^/hr, range 0–2651 m^3^/hr).

**Figure 5:**
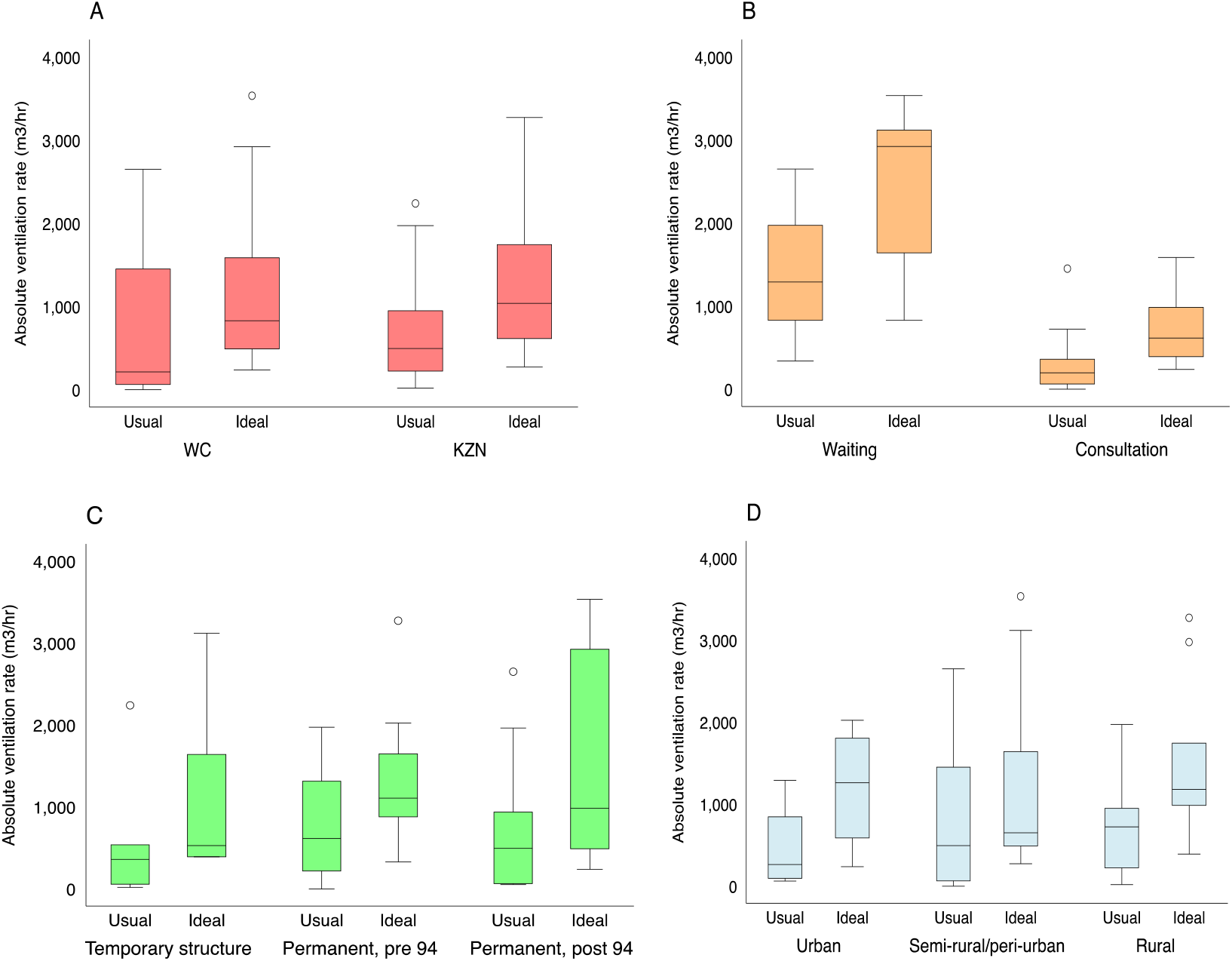
Box and whisker plots describing the distribution of the absolute ventilation rate in the 26 clinical spaces where we conducted tracer gas release experiments. Ventilation rates are described under usual conditions and with all doors and windows fully open. Results are disaggregated by province (A), room type (B), the age of the building (C), and clinic location (D). Here, the boxes mark the 25^th^ percentile, the 50^th^ percentile (median) and the 75^th^ percentile, with the whiskers marking the upper and lower adjacent values.

Supplementary table 3 contains full results for each space in which tracer gas release experiments were performed. Supplementary figure 3 shows the association between wind speed and the absolute ventilation rate for the 26 spaces in which we undertook tracer gas release experiments.

### Rebreathed fraction experiments

The estimated absolute ventilation rates in the eight clinic waiting rooms where the rebreathed fraction approach was used are presented in Figure 6 and supplementary table 4. The volume of these waiting rooms was a median 342.7 (range 50.2–2147.3) m^3^. Under usual conditions, there were a median 4.5 ACH (range 2.1–33.5), which improved under ideal conditions to a median 11.2 ACH [range 2.5–107.2]. Absolute ventilation rates were lower under usual conditions than under ideal conditions (median 1898.8 [range 701.5–4815.1] m^3^/hr vs. 5417.8 [range 1870.3–9328.4] m^3^/hr, respectively) with fully opening existing doors and windows improving the absolute ventilation rate by a median 2.0 fold (range 1.0–6.5).

**Figure 6.**
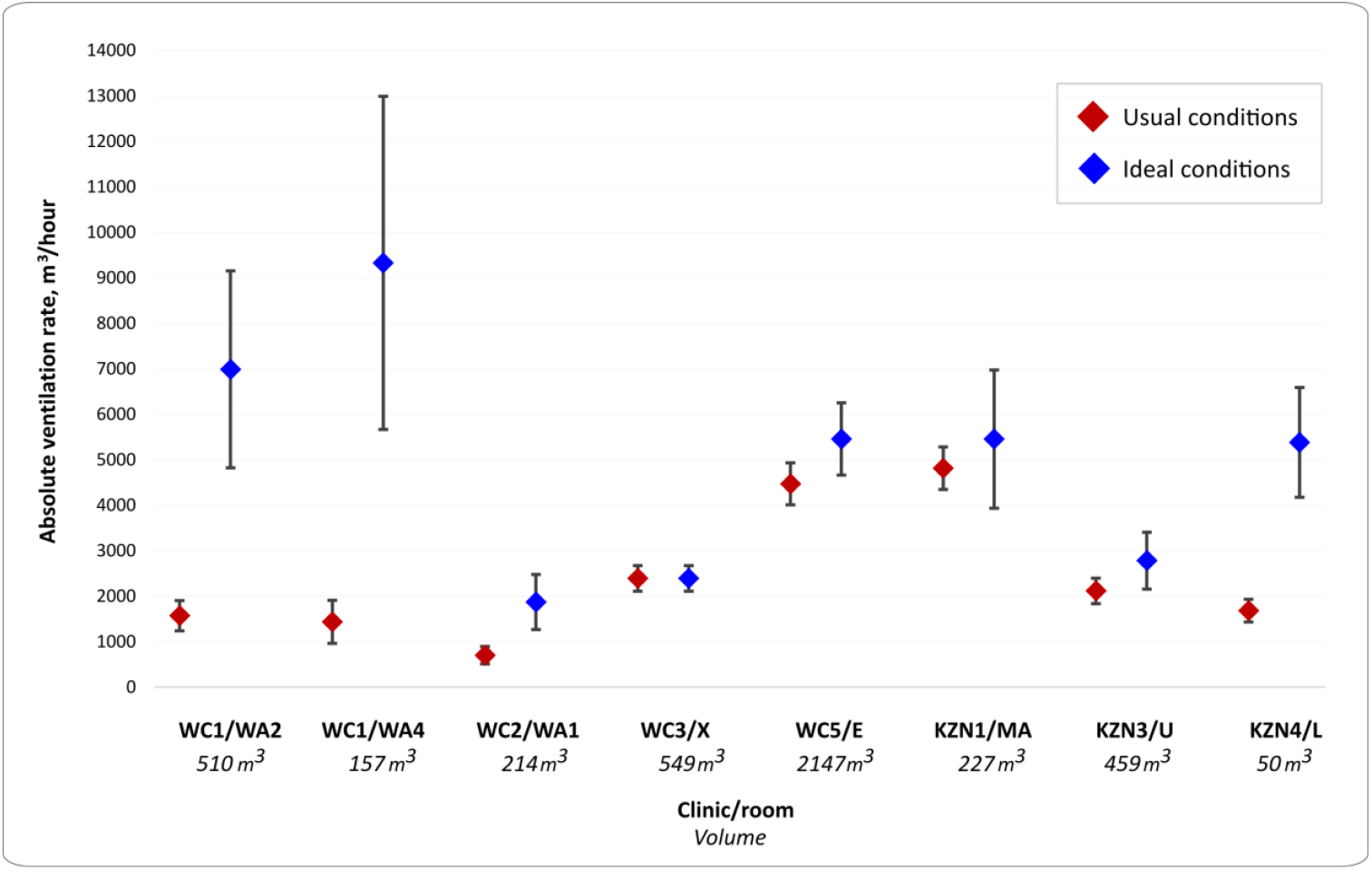
Absolute ventilation rates of eight clinic waiting rooms under usual and ideal conditions, estimated by the rebreathed fraction approach Usual conditions = configuration of windows and doors observed when the room was in routine use Ideal conditions = all windows and doors maximally open Clinic WC3, room X: usual conditions the same as ideal conditions Vertical bars indicate upper and lower estimates of 95% confidence intervals WC: Western Cape; KZN: KwaZulu-Natal; WA2, WA4, WA1, X, E, MA, U, L are the codes for the specific waiting rooms where experiments were performed.

### Estimated risk of *Mycobacterium tuberculosis* transmission

We related the absolute ventilation rate to the risk of Mtb transmission using the Wells Riley Equation[26], as described above. We produced estimates of infection risk under various assumptions about the quanta production rate (Figure 7). The horizontal lines on the figure give the median clinic visit duration of 2 hours 36 minutes [30]. The vertical lines give the median absolute ventilation rate in waiting rooms under both usual and ideal conditions. Combining data from both the tracer gas release experiments and the rebreathed fraction approach, this figure is 1769 m^3^/hr under usual conditions and 2950 m^3^/hr under ideal conditions. Meaningful reductions in transmission risk can be achieved by improving ventilation or reducing the duration of clinic visits. However, ventilation alone cannot eliminate transmission risk in the context of sustained exposure.

**Figure 7.**
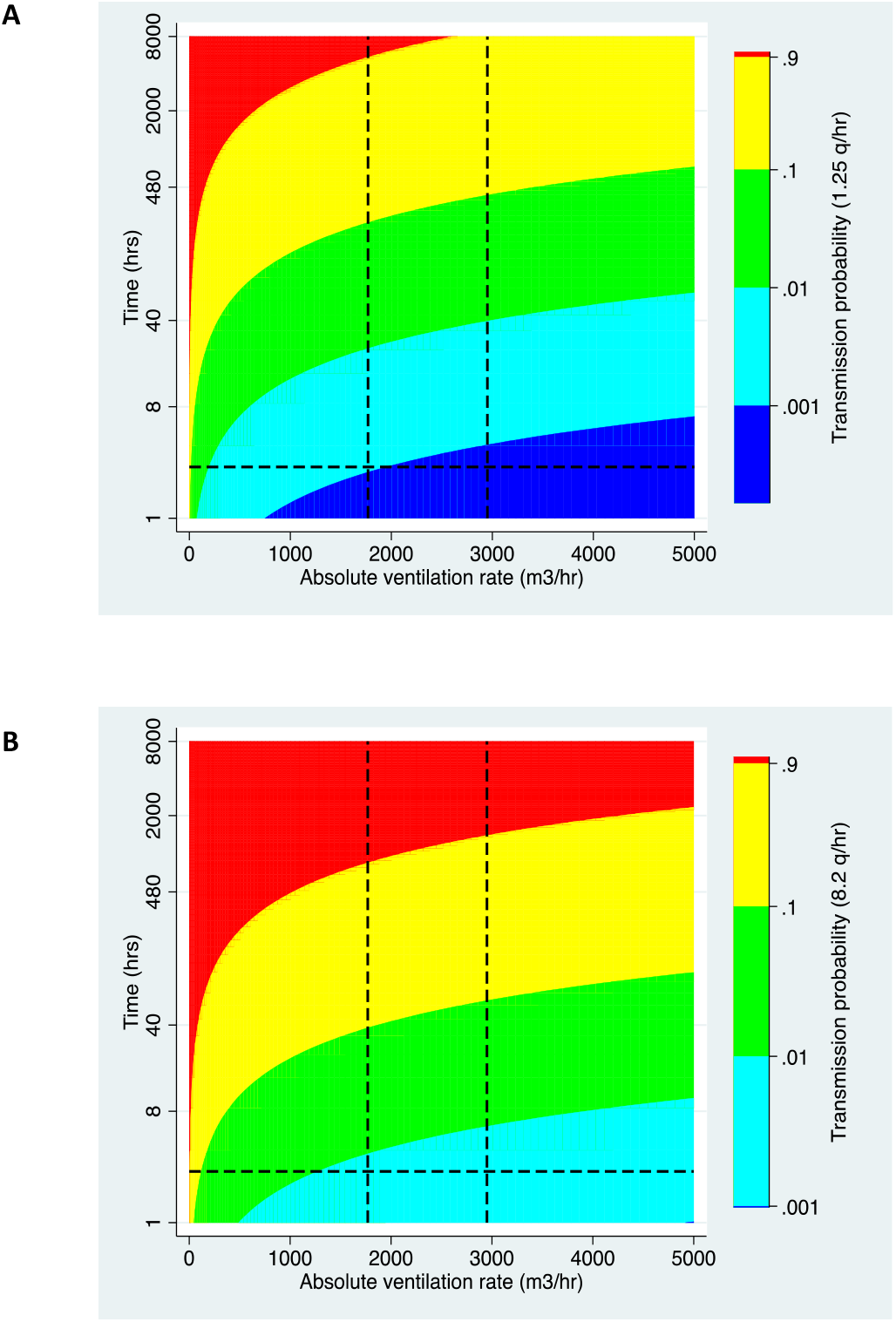
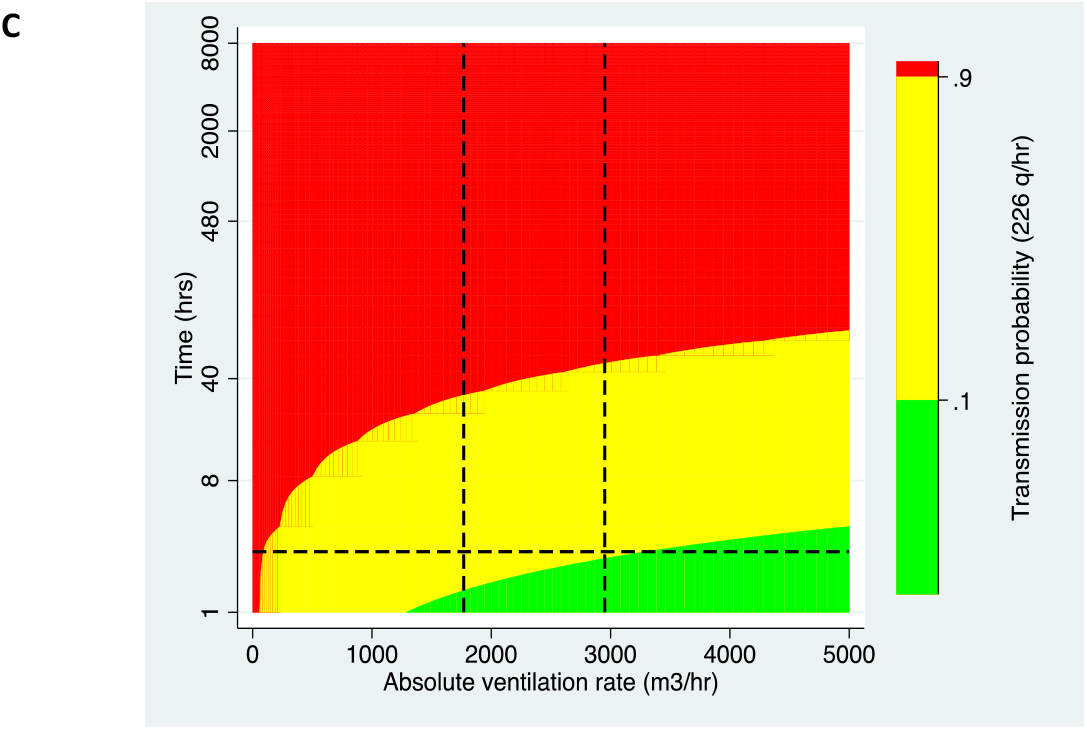
A set of heat maps translating ventilation rate into transmission risk, as estimated using the Wells-Riley equation at 1.25 (A), 8.2 (B) and 226 (C) quanta/hr. The horizontal lines show the median clinic visit duration. The vertical lines show the median absolute ventilation rate in clinic waiting rooms under both usual and ideal conditions.

## Discussion

We observed substantial variation between spaces in rates of natural ventilation. Under usual conditions, using the gold standard tracer gas release method, the median number of ACH was 11.7 and the median absolute ventilation rate was 447 m^3^/hr. There were higher ventilation rates in waiting rooms (median Q = 1289 m^3^/hr) than in consultation rooms (median Q = 197 m^3^/hr). In the eight waiting rooms where we used the rebreathed fraction approach, the median absolute ventilation rate was 1898.8 m^3^/hr. We showed that ventilation rates could, on average, be doubled if existing doors and windows were opened.

There are few published estimates of the absolute ventilation rate in naturally ventilated public spaces, including clinical spaces, in high burden settings. In Peruvian hospitals, Escombe and colleagues estimated an absolute ventilation rate of 2477 m^3^/hr in naturally ventilated spaces, and 402 m^3^/hr in mechanically ventilated spaces [17]. In a study of four small clinic rooms in Cape Town, Cox and colleagues estimated a median ventilation rate of 487 m^3^/hr in the best ventilated space, but none of these experiments were conducted with both the window and the wind-driven roof turbines open [5].

The latest World Health Organization (WHO) guidelines on TB IPC, published in 2019, do not contain a recommended target ventilation rate for clinical spaces [3]. A 2009 WHO guideline recommended that ‘general wards and outpatient spaces’ in naturally ventilated buildings should aim for a minimum ventilation rate of 60 litres/second/person (216 m^3^/hr/person)[1]. For ‘airborne precaution rooms’ (small rooms for accommodating people who may be infectious) the recommended ventilation rate was 160 litres/second/person on average (576 m^3^/hr/person), with a minimum rate of 80 litres/second/person (288 m^3^/hr/person) when wind speed and direction are not favourable. In high burden settings, the transmission risk in general clinic spaces may be equivalent to that in ‘airborne precaution rooms’, given the high prevalence of undiagnosed TB [31]. Dividing the ventilation rates estimated in this study by the per person rates recommended by the WHO yields estimates of maximum occupancy that are lower than those we observed in these clinics. The median ventilation rate in waiting rooms (combining both measurement approaches) meant only eight people could be accommodated under usual conditions, or 13 under ideal conditions, whilst achieving a ventilation rate of 60 litres/second/person. The median ventilation rate in consultation rooms meant doors and windows needed to be fully opened for a health worker and one patient to be accommodated whilst achieving a ventilation rate of 60 litres/second/person. We undertook detailed analysis of patient flow in nine waiting rooms at three clinics, as part of the *Umoya omuhle* project. The quietest waiting area, in a rural clinic, had a median occupancy of 11 people, whereas the busiest waiting area had a median occupancy of 52 people [30]. Consultation rooms typically accommodate a single healthcare worker plus a patient, though patients are not infrequently accompanied by a friend or family member. The fold changes in ventilation rates needed to meet these WHO recommendations may be unfeasible without significant decongestion of clinic spaces.

We showed that natural ventilation can be improved by two-fold by fully opening available windows and doors. We observed that only 30% of spaces had their windows fully open and all consulting room doors were closed when consultations were taking place. Major barriers to opening existing doors and windows include concerns regards privacy and thermal comfort [32]. In a number of spaces, we observed doors and windows had been closed to allow air conditioning units to function. Detailed qualitative research exploring TB IPC related beliefs and behaviours in the same clinics will be presented separately.

An advantage of this work is the wide range of clinical spaces studied. These included: both new and old buildings, temporary and permanent structures, in urban and rural clinics across two provinces. We found that small rooms in temporary structures were particularly badly ventilated. Investment in replacing temporary structures with well-designed permanent structures should be considered, including, where feasible, covered outdoor waiting areas. Low-cost adaptations to existing structures should also be considered. A study of small, poorly-ventilated clinic rooms in Cape Town demonstrated improvements in natural ventilation associated with use of wind driven roof turbines or ‘Whirlybird’ fans [5]. A study in Lima, Peru measured ventilation before and after making changes to six clinical spaces. These changes ranged in intensity from repairing windows that could not be opened at a cost of USD 25, to building a sheltered outdoor waiting area at a cost of USD 7000. The changes resulted in a median 3.0 fold increase in the ventilation rate [33].

Our Wells-Riley estimates of *Mtb* transmission risk suggest that substantial reductions in transmission risk can be achieved by improvements in both natural ventilation and the duration of clinic visits. The substantial variation in ventilation rates observed among spaces within clinics also suggest that transmission risk might be reduced by reorganising care so that patients spend more time in better ventilated spaces. However, as has been argued elsewhere [34], in the absence of other precautions (e.g. consistent use of N95 respirators), very high ventilation rates are needed to protect individuals with prolonged exposure, such as healthcare workers. Where feasible, this might be achieved by moving waiting areas outdoors. The use of upper room germicidal ultraviolet (UVGI) has been advocated where there are constraints on the degree of natural ventilation that can be achieved [32]. This technology is effective at reducing *Mtb* transmission [35][36], and can offer protection on calm days and with doors and windows closed. However, recent experience in South Africa of low-quality installations and poor maintenance led to it falling out of favour [37]. UVGI implementation in resource limited settings is an active area of research and innovative approaches may facilitate easier installation and maintenance [38].

As part of the wider *Umoya omuhle* project, we have modelled the impact of IPC interventions implemented in clinics, including improvements in natural ventilation, on transmission both within clinics [39] and in surrounding communities [40]. Cost effectiveness analyses of these interventions will be published shortly. This work considers the infrastructural, organisational and behavioural changes needed for such interventions to be sustainable. These results support additional investment in a package of IPC interventions to limit the transmission of *Mtb* and other airborne pathogens. Ideally, these interventions would be introduced in manner that allowed robust estimation of their impact, as has previously been done during the national roll out of major interventions in South African healthcare system [41].

CO_2_ release experiments are a robust approach to measuring ventilation. However, the approach is labour intensive, technically demanding, and not possible in large open spaces or spaces that cannot be emptied of people. For this reason, we obtained ventilation estimates using this approach in a limited number of spaces and, usually, only on a single visit. As such, we will not have captured the variability in ventilation that might be expected with changes in weather and season. However, considering these spaces, we obtained measurements over the course of a calendar year. The days on which we took our measurements were broadly representative with respect to temperature and wind speed. Mathematically, the rebreathed fraction approach should give comparable results, though it required assumptions about occupants’ metabolic rate. Direct comparison between the two approaches, in the same space and with the same weather conditions, was not possible, because one method required the space to be occupied and other required it to be empty. It should be noted that while the confidence intervals around the estimates obtained using the rebreathed fraction approach were frequently large, this reflects changes in ventilation rates that occurred over the measurement period – for instance, due to changes in wind speed – as well as the precision of our measurements.

Simpler approaches to estimating the absolute ventilation rate are needed. Several have been proposed, such as simply measuring indoor CO_2_ levels [42], or estimating transmission risk using the approach described by Rudnick and Milton [23]. However, these approaches do not partition risk into that caused by poor ventilation versus that caused by overcrowding, problems with distinct solutions. A single infectious person may transmit to a single susceptible person in a poorly ventilated space with the low room occupancy only resulting in modest rises in background CO_2_ levels. Furthermore, in our experience, rebreathed fraction measurements may be difficult to interpret where there is rapid flux in levels of occupancy – i.e. in small consultation rooms where patients enter and leave frequently.

Ideally, we need approaches to estimate the absolute ventilation rate that do not depend on costly equipment; that can be performed quickly by clinic staff in occupied spaces with minimal training; and with immediate results to guide risk reduction interventions, such as reducing occupancy or opening windows. A possible approach advocated in WHO documents [1][2] makes a set of simple calculations based on wind speed, which must be measured, and the area of the windows of windows and doors. No validation data are presented and the method can only be applied in spaces with openings on opposite walls. One alternative might be a simplified tracer gas release experiment, ideally using an inert substance not usually present in room air. This might mean that small volumes of tracer gas could be released with the room in routine use. We wonder whether concentrations might then be measured using a meter adapted to feed data to a smart phone, with a phone application used to interpret the decay curve.

In conclusion, we observed substantial variation in natural ventilation in clinical spaces in primary health care facilities in two provinces of South Africa. The worst ventilated spaces were small rooms where doors and windows had been closed, and temporary structures had lower ventilation rates than permanent structures. Opening all existing doors and windows resulted in meaningful improvements in ventilation. Concerns regarding privacy and thermal comfort may place limits on the ventilation rates that can be achieved. A package of IPC interventions, including those directed at improving natural ventilation, are needed to reduce *Mtb* transmission in these settings. In future work, we plan to model adaptations to clinic structures that maximise ventilation without compromising thermal comfort.

## Data Availability

This Stata code for the analysis of the tracer gas release experiments is available on GitHub
(https://github.com/tayates/uo_ventilation). Analysis of paired indoor-outdoor CO2 measurements
was carried out using R version 3.6.0 [29].This R code is also available on GitHub (URL:https://github.com/ArminderD/ventilation.git). The data are available via https://datacompass.lshtm.ac.uk/.

## Acknowledgments

We are grateful to the clinical and management staff at 10 clinics where we obtained ventilation measurements. We thank Thomas Murray, Harriet Gliddon, and Sinethemba Mabuyakhulu who assisted us with ventilation measurements in KZN. We are grateful to Rod Escombe, Ed Nardell, Jon Taylor, Don Milton and Toby van Reenen for useful discussions about various aspects of ventilation science – they take no responsibility for the content of this manuscript.

## Author contributions

Conceptualization:TAY, ADG, PGB

Data collection and curation: PGB, ASK, IG

Formal analysis: TAY, AD, PGB, ASK, KB

Funding acquisition: ADG, NM, TAY, KB, KK

Methodology: PGB, ASK, TAY, AD, NM, KB, ADG

Project administration: ASK, TAY, ADG, KK

Visualization: PGB, ASK, TAY

Writing – original draft: PGB

Writing – review & editing: ASK, IG, AD, NM, KB, KK, ADG, TAY

## Supplementary material

Supplementary table 1: Differences between the two methods used to measure ventilation in the clinics

Supplementary table 2: Definitions of usual and ideal conditions

Supplementary table 3: Absolute ventilation rates of the clinic spaces under usual and ideal conditions using the tracer gas release technique

Supplementary table 4: Ventilation rates using the rebreathed fraction approach using the continuous measurements under usual and ideal conditions

Supplementary Figure 1: an example of a tracer gas release experiment from Room C at KZN2 clinic (experiment 6 Monitor a). Following log transformation, the right hand side of the curve is approximately linear.

Supplementary Figure 2: Histograms showing the distribution of temperatures and wind speeds in KwaZulu-Natal and Western Cape during working hours from January 2018 – December 2020. Vertical lines shows the mean temperatures and wind speeds on the 8 days when the rebreathed approach experiments were conducted.

Supplementary Figure 3: Association between wind speed and the absolute ventilation rates

## Supplementary material

### Supplementary Tables

**Supplementary table 1:** Differences between the two methods used to measure ventilation in the clinics

| Name of method | Brief description of method | Room conditions in which experiments performed | Occupancy of space at time of experiment | Rooms in which experiments were performed in | Advantages | Disadvantages |
| --- | --- | --- | --- | --- | --- | --- |
| Tracer gas release experiments | CO<sub>2</sub> was used as the tracer gas for this method. CO<sub>2</sub> was released. The windows and doors were opened and the decay of the CO<sub>2</sub> was measured. | Ideal and usual conditions | Rooms had no HCWs or patients in the room | Mainly small spaces where a high CO<sub>2</sub> baseline could be achieved | Gold standard | CO<sub>2</sub> is released into the atmosphere<br>Difficult to measure large spaces with this method<br>Difficult to perform during work hours in areas with a large occupancy |
| Rebreathed fraction approach | CO<sub>2</sub> levels were measured every second in three inside sites of the waiting room and at one site outside of the building. Headcounts were performed. | Ideal and usual conditions | Rooms were occupied by HCWs and patients | Main waiting rooms | Can be performed during working hours. | Headcounts were performed every 10 minutes. The waiting rooms were very dynamic so the variations of CO<sub>2</sub> produced by the number of people may not have been captured |

CO<sub>2</sub>: Carbon dioxide HCW: healthcare workers

**Supplementary table 2:** Definitions of Usual and Ideal conditions

| **Term** | **Description** |
| --- | --- |
| Usual conditions | The configuration of the existing windows and doors in the room observed on the day of room volume measurements (in the case of the tracer gas release experiments); or<br>The configuration of the existing windows and doors found when in use (in the case of the rebreathed fraction approach) |
| Ideal conditions | All windows and doors fully opened to allow for maximal ventilation |

**Supplementary table 3:**
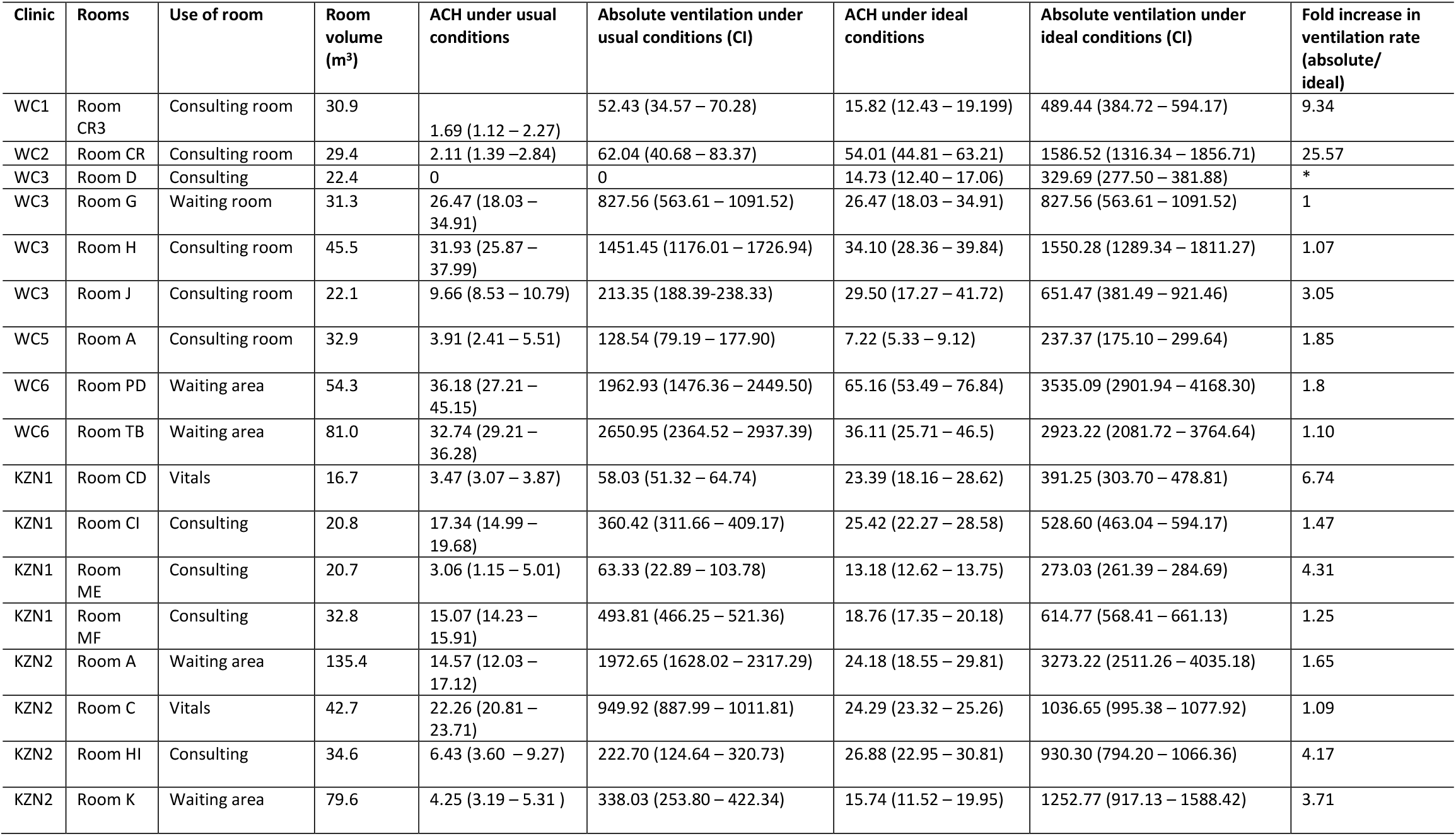
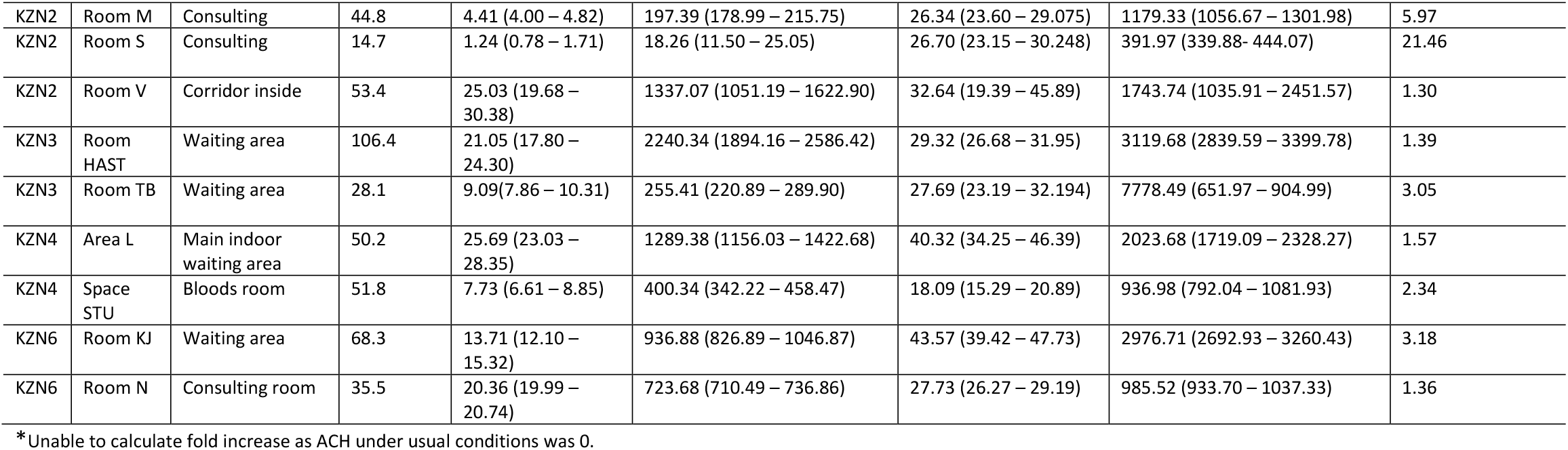
Absolute ventilation rates of the clinic spaces under usual and ideal conditions using the tracer gas release technique

| Clinic | Rooms | Use of room | Room volume (m³) | ACH under usual conditions | Absolute ventilation under usual conditions (CI) | ACH under ideal conditions | Absolute ventilation under ideal conditions (CI) | Fold increase in ventilation rate (absolute/ideal) |
| --- | --- | --- | --- | --- | --- | --- | --- | --- |
| WC1 | Room CR3 | Consulting room | 30.9 | 1.69 (1.12 – 2.27) | 52.43 (34.57 – 70.28) | 15.82 (12.43 – 19.199) | 489.44 (384.72 – 594.17) | 9.34 |
| WC2 | Room CR | Consulting room | 29.4 | 2.11 (1.39 – 2.84) | 62.04 (40.68 – 83.37) | 54.01 (44.81 – 63.21) | 1586.52 (1316.34 – 1856.71) | 25.57 |
| WC3 | Room D | Consulting | 22.4 | 0 | 0 | 14.73 (12.40 – 17.06) | 329.69 (277.50 – 381.88) | * |
| WC3 | Room G | Waiting room | 31.3 | 26.47 (18.03 – 34.91) | 827.56 (563.61 – 1091.52) | 26.47 (18.03 – 34.91) | 827.56 (563.61 – 1091.52) | 1 |
| WC3 | Room H | Consulting room | 45.5 | 31.93 (25.87 – 37.99) | 1451.45 (1176.01 – 1726.94) | 34.10 (28.36 – 39.84) | 1550.28 (1289.34 – 1811.27) | 1.07 |
| WC3 | Room J | Consulting room | 22.1 | 9.66 (8.53 – 10.79) | 213.35 (188.39–238.33) | 29.50 (17.27 – 41.72) | 651.47 (381.49 – 921.46) | 3.05 |
| WC5 | Room A | Consulting room | 32.9 | 3.91 (2.41 – 5.51) | 128.54 (79.19 – 177.90) | 7.22 (5.33 – 9.12) | 237.37 (175.10 – 299.64) | 1.85 |
| WC6 | Room PD | Waiting area | 54.3 | 36.18 (27.21 – 45.15) | 1962.93 (1476.36 – 2449.50) | 65.16 (53.49 – 76.84) | 3535.09 (2901.94 – 4168.30) | 1.8 |
| WC6 | Room TB | Waiting area | 81.0 | 32.74 (29.21 – 36.28) | 2650.95 (2364.52 – 2937.39) | 36.11 (25.71 – 46.5) | 2923.22 (2081.72 – 3764.64) | 1.10 |
| KZN1 | Room CD | Vitals | 16.7 | 3.47 (3.07 – 3.87) | 58.03 (51.32 – 64.74) | 23.39 (18.16 – 28.62) | 391.25 (303.70 – 478.81) | 6.74 |
| KZN1 | Room CI | Consulting | 20.8 | 17.34 (14.99 – 19.68) | 360.42 (311.66 – 409.17) | 25.42 (22.27 – 28.58) | 528.60 (463.04 – 594.17) | 1.47 |
| KZN1 | Room ME | Consulting | 20.7 | 3.06 (1.15 – 5.01) | 63.33 (22.89 – 103.78) | 13.18 (12.62 – 13.75) | 273.03 (261.39 – 284.69) | 4.31 |
| KZN1 | Room MF | Consulting | 32.8 | 15.07 (14.23 – 15.91) | 493.81 (466.25 – 521.36) | 18.76 (17.35 – 20.18) | 614.77 (568.41 – 661.13) | 1.25 |
| KZN2 | Room A | Waiting area | 135.4 | 14.57 (12.03 – 17.12) | 1972.65 (1628.02 – 2317.29) | 24.18 (18.55 – 29.81) | 3273.22 (2511.26 – 4035.18) | 1.65 |
| KZN2 | Room C | Vitals | 42.7 | 22.26 (20.81 – 23.71) | 949.92 (887.99 – 1011.81) | 24.29 (23.32 – 25.26) | 1036.65 (995.38 – 1077.92) | 1.09 |
| KZN2 | Room HI | Consulting | 34.6 | 6.43 (3.60 – 9.27) | 222.70 (124.64 – 320.73) | 26.88 (22.95 – 30.81) | 930.30 (794.20 – 1066.36) | 4.17 |
| KZN2 | Room K | Waiting area | 79.6 | 4.25 (3.19 – 5.31 ) | 338.03 (253.80 – 422.34) | 15.74 (11.52 – 19.95) | 1252.77 (917.13 – 1588.42) | 3.71 |
| KZN2 | Room M | Consulting | 44.8 | 4.41 (4.00 – 4.82) | 197.39 (178.99 – 215.75) | 26.34 (23.60 – 29.075) | 1179.33 (1056.67 – 1301.98) | 5.97 |
| KZN2 | Room S | Consulting | 14.7 | 1.24 (0.78 – 1.71) | 18.26 (11.50 – 25.05) | 26.70 (23.15 – 30.248) | 391.97 (339.88- 444.07) | 21.46 |
| KZN2 | Room V | Corridor inside | 53.4 | 25.03 (19.68 – 30.38) | 1337.07 (1051.19 – 1622.90) | 32.64 (19.39 – 45.89) | 1743.74 (1035.91 – 2451.57) | 1.30 |
| KZN3 | Room HAST | Waiting area | 106.4 | 21.05 (17.80 – 24.30) | 2240.34 (1894.16 – 2586.42) | 29.32 (26.68 – 31.95) | 3119.68 (2839.59 – 3399.78) | 1.39 |
| KZN3 | Room TB | Waiting area | 28.1 | 9.09(7.86 – 10.31) | 255.41 (220.89 – 289.90) | 27.69 (23.19 – 32.194) | 7778.49 (651.97 – 904.99) | 3.05 |
| KZN4 | Area L | Main indoor waiting area | 50.2 | 25.69 (23.03 – 28.35) | 1289.38 (1156.03 – 1422.68) | 40.32 (34.25 – 46.39) | 2023.68 (1719.09 – 2328.27) | 1.57 |
| KZN4 | Space STU | Bloods room | 51.8 | 7.73 (6.61 – 8.85) | 400.34 (342.22 – 458.47) | 18.09 (15.29 – 20.89) | 936.98 (792.04 – 1081.93) | 2.34 |
| KZN6 | Room KJ | Waiting area | 68.3 | 13.71 (12.10 – 15.32) | 936.88 (826.89 – 1046.87) | 43.57 (39.42 – 47.73) | 2976.71 (2692.93 – 3260.43) | 3.18 |
| KZN6 | Room N | Consulting room | 35.5 | 20.36 (19.99 – 20.74) | 723.68 (710.49 – 736.86) | 27.73 (26.27 – 29.19) | 985.52 (933.70 – 1037.33) | 1.36 |

\*Unable to calculate fold increase as ACH under usual conditions was 0.

**Supplementary table 4:** Ventilation rates using the rebreathed fraction approach using continuous measurements under usual and ideal conditions

| Facility | Rooms | Volume of room | ACH under usual conditions (95% CI) | Absolute ventilation under usual conditions(95% CI) l/s | Absolute ventilation under usual conditions(95% C/l) m<sup>3</sup>/hr | ACH under ideal conditions (95% CI) | Absolute ventilation under ideal conditions (95% CI) l/s | Absolute ventilation under ideal conditions(95% C/l) m<sup>3</sup>/hr | Fold increase in ventilation |
| --- | --- | --- | --- | --- | --- | --- | --- | --- | --- |
| Facility WC1 | Room WA2 | 509.6 | 3.09<br>(2.43 – 3.74) | 436.71<br>(344.08 – 529.34) | 1572.16 (1238.69 – 1905.62) | 13.71<br>(9.46 – 17.96) | 1940.78<br>(1339.33 - 2542.23) | 6986.81 (4821.59 – 9152.03) | 4.44 |
|  | Room WA4 | 157.3 | 9.13<br>(6.12 – 12.14) | 399.14<br>(267.61 – 530.68) | 1436.90 (963.40 – 1910.45) | 59.29<br>(36.01 – 82.57) | 2591.21<br>(1573.8 - 3608.62) | 9328.36 (5665.68 – 12991.03) | 6.49 |
| Facility WC2 | Room WA1 | 213.9 | 3.28<br>(2.37 – 4.19) | 194.85<br>(140.96 – 248.73) | 701.46 (507.456 – 895.43) | 8.74<br>(5.91 – 11.58) | 519.53<br>(351.24 - 687.82) | 1870.31 (1264.46 – 2476.15) | 2.66 |
| Facility WC3 | Room X | 549.0 | 4.36<br>(3.84 – 4.87) | 664.3<br>(586.37 – 742.23) | 2391.48 (2210.93 – 2672.03) | 4.36<br>(3.84 – 4.87) | 664.3<br>(586.37 - 742.23) | 2391.48 (2210.93 – 2672.03) | 1 |
| Facility WC5 | Room E | 2147.3 | 2.08<br>(1.87 – 2.3) | 1241.75<br>(1113.2 – 1370.29) | 4470.3 (4007.52 – 4933.04) | 2.54<br>(2.17 – 2.91) | 1516.34<br>(1295.59 - 1737.09) | 5458.82 (4664.12 – 6253.52) | 1.22 |
| Facility KZN1 | Room MA | 226.7 | 21.24<br>(19.18 – 23.3) | 1337.53<br>(1207.89 – 1467.18) | 4815.11 (4348.40 – 5281.85) | 24.06<br>(17.35 – 30.78) | 1515.17<br>(1092.53 - 1937.82) | 5454.61 (3933.11 – 6976.15) | 1.13 |
| Facility KZN3 | Room U | 458.7 | 4.62<br>(4.00 – 5.23) | 588.04<br>(509.63 – 666.44) | 2116.94 (1834.67 – 2399.18) | 6.07<br>(4.7 – 7.43) | 772.79<br>(598.84 - 946.75) | 2782.04 (2155.82 – 3408.3) | 1.31 |
| Facility KZN4 | Space L | 50.2 | 33.49<br>(28.51 – 38.46) | 466.83<br>(397.52 – 536.14) | 1680.59 (1431.07 – 1930.10) | 107.22<br>(83.15 – 131.29) | 1494.74<br>(1159.15 - 1830.33) | 5381.06 (4172.94 – 6589.19) | 3.20 |

### Figures

**Supplementary Figure 1.**
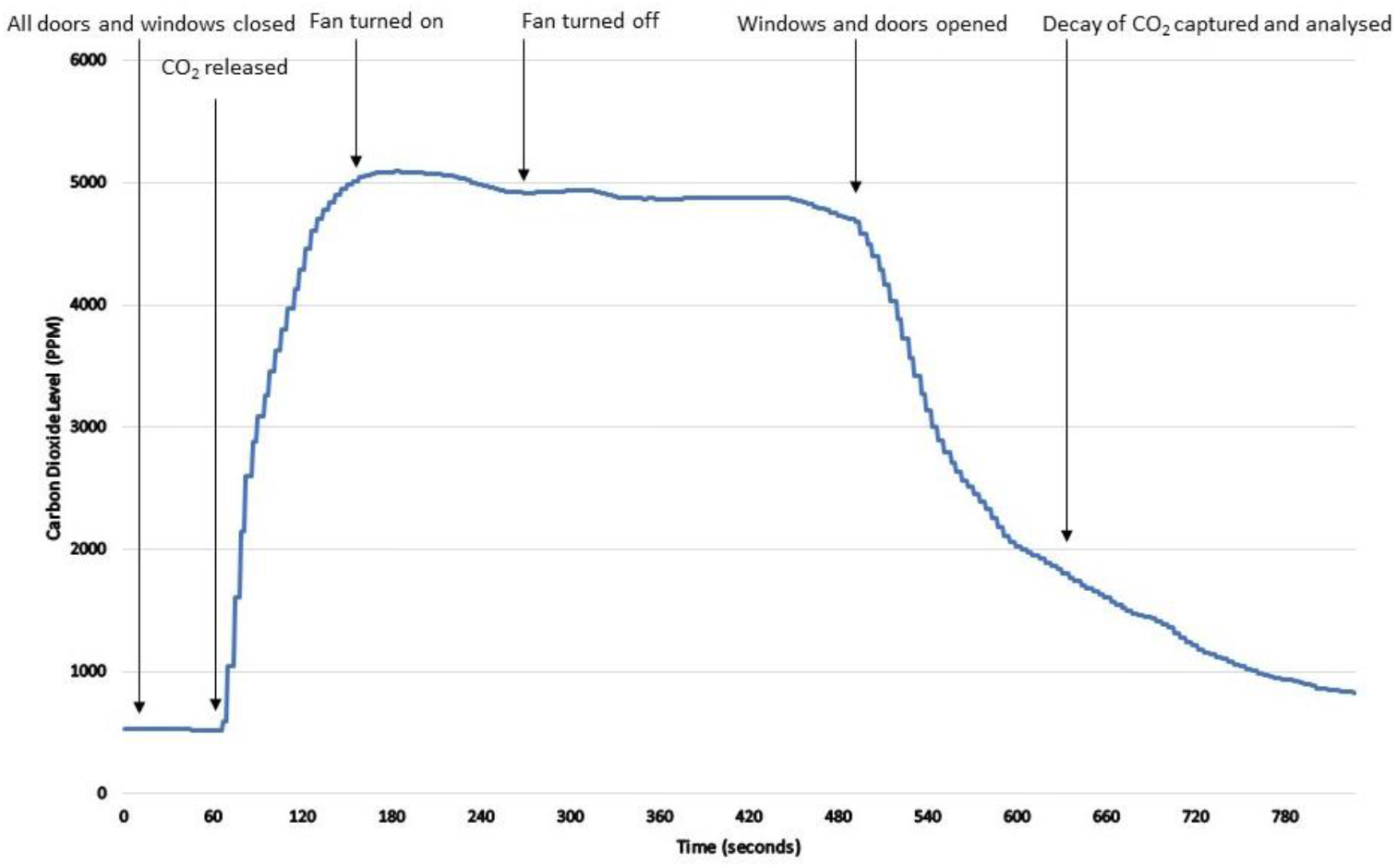
*An example of a tracer gas release experiment from Room C at KZN2 clinic (experiment 6 Monitor A). Following log transformation, the right hand side of the curve is approximately linear*. CO_2_: carbon dioxide; PPM: parts per million

**Supplementary Figure 2:**
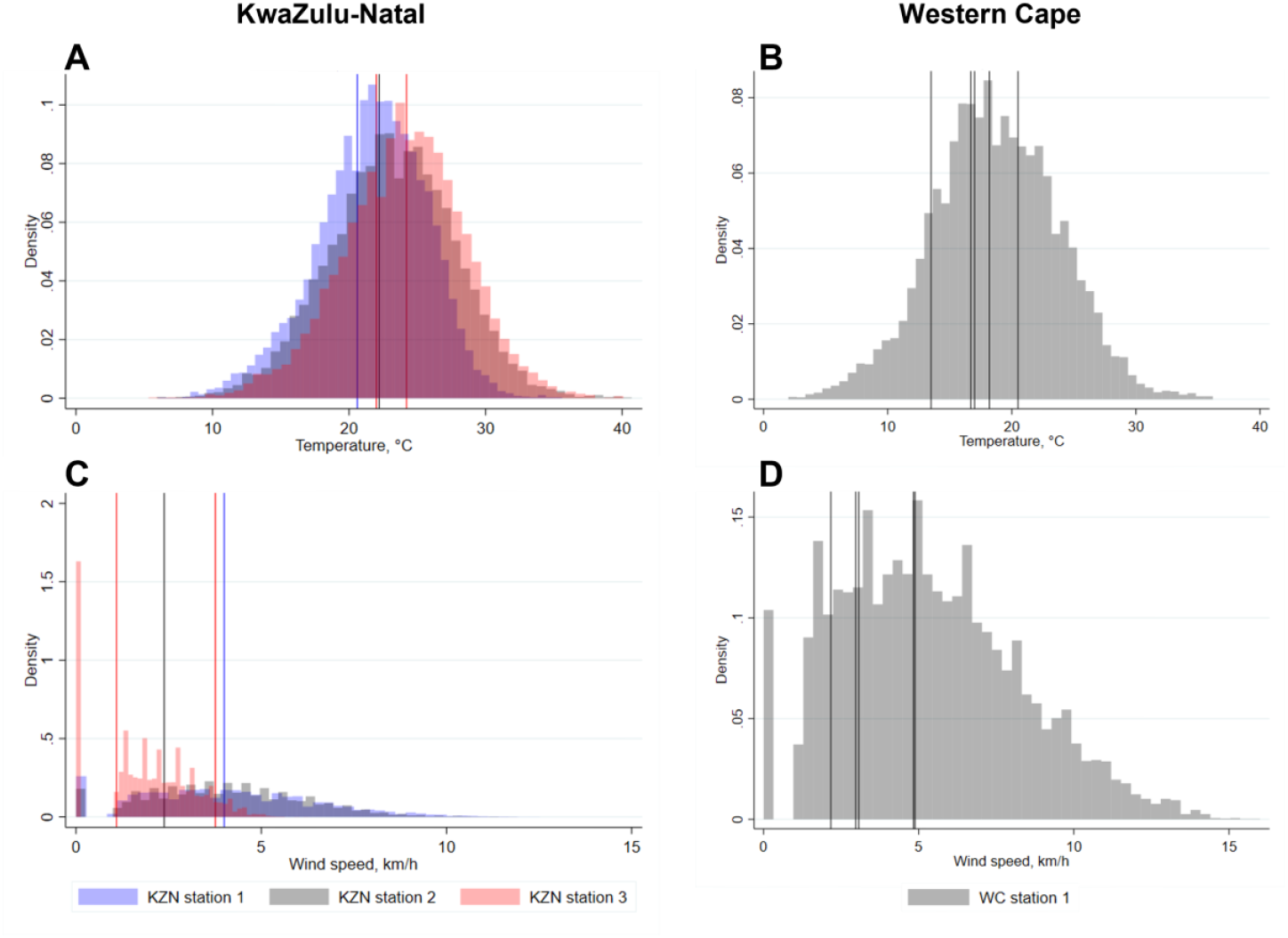
Histograms showing the distribution of temperatures and wind speeds in KwaZulu-Natal and Western Cape during working hours from January 2018 - December 2020. Vertical lines shows the mean temperatures and wind speeds on the 8 days when the rebreathed approach experiments were conducted

**Supplementary Figure 3:**
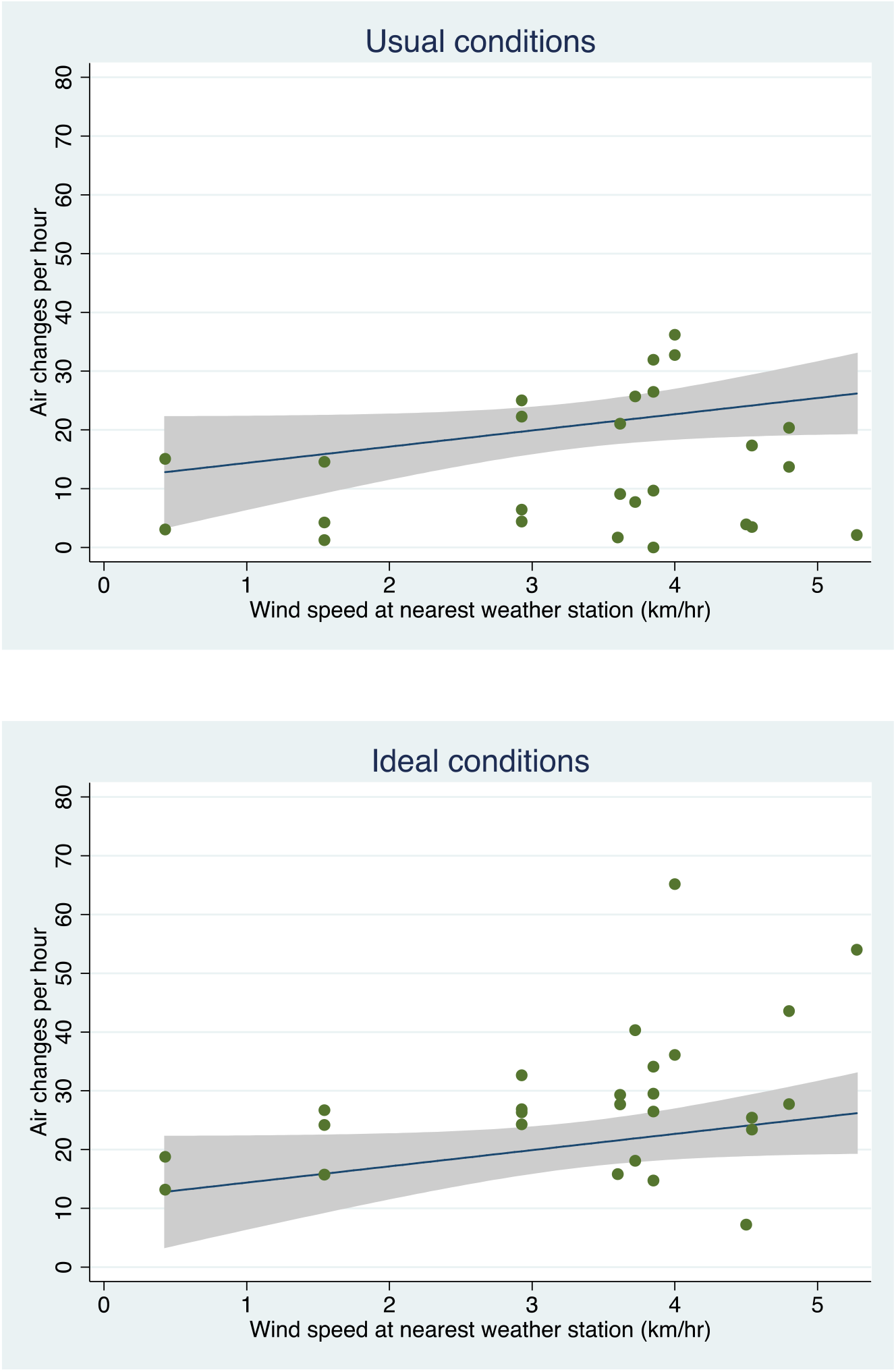
The association between wind speed at the nearest weather station and the number of air changes per hour in 26 clinical spaces. Measurements were taken both under usual conditions (Beta: 1.56, 95% CI [-1.98 – 5.10]; R2: 0.03) and ideal conditions (Beta: 3.96, 95 CI [0.23 – 7.70]; R2: 0.17)

